# Ethical Considerations during the Informed Consent Process for Acute Ischemic Stroke in International Clinical Trials

**DOI:** 10.1101/2020.10.14.20212779

**Authors:** Tiffany R. Bellomo, Jennifer A. Fokas, Clare Anderson, Noah Tsao, Christopher Becker, Rachel Gioscia-Ryan, William J. Meurer

**Author notes:** Authors contributed equally to this work (Tiffany R. Bellomo, Jennifer A. Fokas). Corresponding Author:* William J. Meurer, MD, Department of Emergency Medicine, University of Michigan Medical School, 1500 E Medical Center Drive, Ann Arbor, MI 48109-5303 USA.

## Abstract

**Introduction:** Obtaining informed consent from acute ischemic stroke patients poses many challenges, especially in the context of a research setting. Specifically, consenting for alternative acute ischemic stroke treatments to the standard of care, Tissue Plasminogen Activator (tPA), can cause delays leading to increased time to reperfusion and worse outcomes.

**Objectives:** We sought to investigate the experiences of researchers in existing active-control trials in acute ischemic stroke comparing investigational therapy to tPA in order to identify the approaches and challenges in obtaining informed consent in this unique patient population.

**Methods:** Out of 401 articles evaluated, 14 trials met inclusion criteria of patients receiving IV tPA vs alternative treatment within 4.5 hours of onset of symptoms for acute ischemic stroke. Trial representatives were emailed by the study team with a request for a copy of their patient consent form, other documents related to informed consent, and to complete a survey concerning aspects of the consent process.

**Results:** Of the 6 trials conducted across 6 continents that completed the survey in its entirety, 2 were ongoing, 4 were published between 2009 and 2016, and the median NIHSS for each published trial was at least an 8. All published trials in the sample stated that informed consent was obtained, but only half reported involvement of a research ethics committee. Although 3 trials performed in Europe or Asia reported directly consenting 75-100% of enrolled patients, the median NIHSS for these trials represented a moderate stroke. Trials with 75-100% of patients directly consented had shorter door to treatment (DTT) times than trials that directly consented less than 50% of enrolled patients. 5 trials allowed consent by proxy, but only 2 of those trials also required patient assent. 4 trials had translators available and translated consent documents, and these trials had longer DTT times. All trials relied on experienced providers or dedicated research coordinators to obtain informed consent; however, only 2 of 6 trials mentioned specific training with regards to informed consent skills.

**Conclusions:** The current informed consent process is not transparent and poses challenges to investigators in the USA directly comparing tPA to an alternative treatment. International differences in the standards of informed consent, such as deferred consent, may have allowed more patients with moderate strokes to provide direct consent after treatment administration without delaying DTT time. While targeted and innovative approaches for informed consent are needed to improve patient outcomes, we must balance protecting the autonomy of individuals whose willing involvement enables such pivotal discoveries. The stroke community must aim for efficiency, transparency, and inclusion of patients of diverse backgrounds in the informed consent process so that therapeutic advances are possible.

## INTRODUCTION

Although the purpose of medical research involving human subjects is to generate knowledge that will potentially benefit a target population, this purpose should not take precedence over the rights of the individual research subjects.(1) Informed consent is the process by which permission is granted for a medical intervention: each human subject is informed of the aims, methods, risks, potential benefits, alternative treatments, and other relevant aspects of the study. The process of informed consent becomes challenging when a study is designed to intervene in an urgent and time-sensitive setting, such as stroke.

The prevalence of stroke in the United States of America (USA) in 2016 was 2.5%, which is projected to increase to 3.9% by 2030.(2,3) Given this increasing burden, a large effort has been made to improve upon current stroke treatments. Administration of IV Tissue Plasminogen Activator (tPA) within 4.5 hours and mechanical thrombectomy for large vessel occlusions within 6 hours of symptoms onset are the standard of care for eligible patients according to the American Heart Association (AHA) guidelines for acute ischemic stroke with significant functional deficits.(4,5) Reperfusion therapies in acute stroke show a clear time-dependent effect, being more effective the earlier treatment is initiated or reperfusion is achieved.(6–8) Therefore, time constraints on informed consent make the consent process difficult and may pose a barrier to examining new and potentially more beneficial therapies.(9–12)

Studying an alternative to tPA involves randomizing patients to tPA vs. investigational therapy; however, the brief informed consent process for administering tPA under high level recommendation alone without the added time burden of a formal consent process for a non-standard-of-care therapy has been reported to delay treatment.(13) In addition, patients with severe stroke symptoms may not have the capacity to consent for research, and thus require consent by proxy (14), which has also been shown to delay treatment, especially when consensus is needed among multiple family members.(13,15) Another source of delay could be elderly patients with cognitive impairment, a population that commonly suffers from stroke. Not only do cognitive impairments complicate determination of capacity, but elderly patients may present to the emergency room unaccompanied, making it difficult to ascertain if they have a surrogate decision maker.(16) Although there has been investigation of alternate methods such as targeted consent models to reduce delays in treatment (17), ethical considerations involved in conducting clinical research in the urgent setting with respect to the process of obtaining informed consent have been an area of ongoing deliberation.(12)

In this study, we investigate the experiences of researchers in existing active-control trials in acute ischemic stroke comparing investigational therapy to tPA in order to identify the approaches and challenges in obtaining informed consent in this unique patient population. To evaluate the impact of research study factors on patient care, we also collected information on door to treatment (DTT) times of both experimental and control arms of trials. This information can serve to not only identify potential barriers and ethical considerations of performing such studies, but also promote new approaches in the acute setting for a more efficient informed consent process that balances being patient centered with the need to develop new interventions.

## METHODS

This research study was reviewed by the University of Michigan Institutional Review Board for Human Subjects Research and determined to be exempt under IRB #HUM00180410.

### Literature Search

Literature searches with the aid of a University of Michigan research librarian were performed using the following platforms: PubMed (https://pubmed.ncbi.nlm.nih.gov/; Bethesda MD US), clinicaltrials.gov (https://clinicaltrials.gov/; US National Library of Medicine), National Institute of Public Health Clinical Trials Search (https://rctportal.niph.go.jp/en/; Japan), Europe PMC (https://europepmc.org/; Europe), EU Clinical Trials Register (https://www.clinicaltrialsregister.eu/ctr-search/search; The Netherlands), the Australian New Zealand Clinical Trials Registry (https://www.anzctr.org.au/; Australia), and International Clinical Trials Registry Platform (apps.who.int/trialsearch/; World Heath Organization). Peer-reviewed scientific articles published in English language were identified using the following search terms: “alteplase” OR “tPA” OR “Activase” and “ischemic stroke” OR “acute ischemic stroke”. Publication dates were restricted to January 1991 through March 2020. The PubMed search was further filtered by selecting Clinical Trials and Humans. The Clinicaltrials.gov search was further filtered by excluding recruitment status listed as suspended, terminated, withdrawn, or unknown. The Australian New Zealand Clinical Trials Registry search was further filtered by selecting “interventional” trial type. Inclusion criteria for trials were studies of patients with acute ischemic stroke who would be eligible for IV tPA administration within 4.5 hours of symptom onset and comparison of IV tPA versus an alternative treatment. Exclusion criteria were trials where IV tPA was combined with an alternative treatment, or where an alternative treatment was compared to a placebo. Three authors assessed all trials independently for eligibility and subsequently collaborated to agree upon the final trials included. Of the 401 articles uploaded to Rayyan (https://rayyan.qcri.org/; Doha, Qatar) for researcher review, 14 trials met inclusion and exclusion criteria (Figure 1).(18–31)

**Figure 1.**
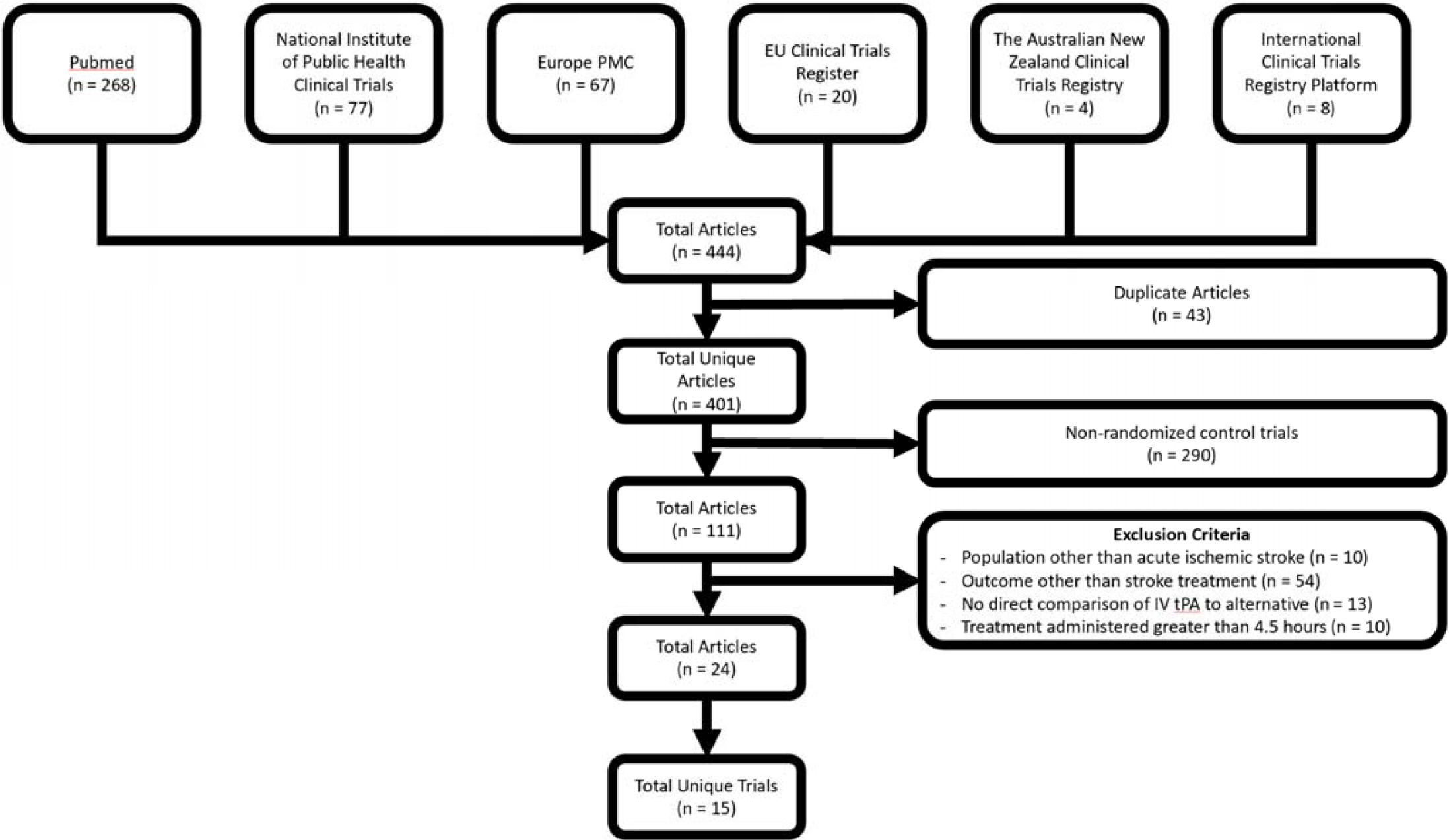
Flowchart of results from the literature search conducted.

### Studies Contacted

The corresponding author and trial coordinators listed on clinicaltrials.gov from each of the 14 trials were emailed by the study team with a request for a copy of their patient consent form, other documents related to informed consent, and to complete a survey developed as described below. The email also included a reminder that participation is voluntary and that emails would be collected at their own discretion. Individuals receiving the survey were asked to share the survey with other members of their trial team who were knowledgeable about informed consent for the trial. The survey contained 24 questions pertaining to the informed consent process, including multiple choice, and free response questions (Supp 1). 43 representatives from the 14 trials were contacted. 6 studies completed the survey in its entirety, 1 study (AcT) partially completed the survey and therefore results were excluded, and the remaining 7 studies did not respond after a third contact.(18–31)

### Survey Development

An anonymous online survey developed using Qualtrics (SAP software company; Utah, USA) assessed how informed consent was obtained in a population that may have challenges with standard consent due to absence of capacity in the setting of acute stroke. Survey questions included demographic questions (year of result publication, location, participants, and median NIHSS of trial arms), multiple choice (yes/no/unknown) questions regarding specific aspects of the informed consent process, and free response questions concerning challenges trials may have faced. A draft of the survey was presented to an acknowledged collaborator, Dr. Lesli Skolarus, where specific feedback was provided on content and format. The survey was then revised and distributed to the 43 contacts mentioned above.

### Statistical Analysis

Informed consent documents from each trial were analyzed for presence or absence of informed consent elements (Table 1). If data was published on median DTT times and the survey respondents used estimates of DTT times, the published DTT times of the trials were used in the statistical analysis. Descriptive statistics were displayed for continuous variables as either mean ± standard deviation or median (interquartile range) depending upon data distributions, and as frequency (percent) for categorical variables. Nonparametric methods (including Wilcoxon Rank Sums and the Kruskal-Wallis test) were used to evaluate factors potentially impacting DTT time. Statistical analyses were performed using R version 3.62 (r-project.org) and Excel (Microsoft Corp; Redmond, Washington, USA).

**Table 1.** Demographic characteristics of the 6 trials that completed the survey, including the median National Institutes of Health Stroke Scale (NIHSS) score, door to treatment times (DTT), and informed consent discussion in the published results from the trial.

|  | **ENCHANTED** | **noR-TEST**<br>**2** | **IV vs. IA**<br>**tPA** | **TNKS2B** | **noR-TEST** | **TPK**<br>**Derivative** |
| --- | --- | --- | --- | --- | --- | --- |
| **Status of Trial** | Completed | Ongoing | Completed | Completed | Completed | Ongoing |
| **Year of Result**<br>**Publication** | 2016 | Ongoing | 2009 | 2010 | 2017 | Ongoing |
| **Trial Location** | Asia, Australia,<br>Europe, South<br>America | Europe | North<br>America | North<br>America | Europe | Asia |
| **Number of**<br>**Participants** | 3206 | Ongoing | 7 | 112 | 1100 | Ongoing |
| **Median NIHSS**<br>**Score (IQR)** | 8 (5-14) | Ongoing | 16 (7-20) | 13 (5-17) | 8 (7-11) | Ongoing |
| **Control Arm**<br>**Median DTT**<br>**(min)** | 170 | 10 | 180 | 65 | 34 | 43 |
| **Experimental**<br>**Arm Median DTT**<br>**(min)** | 170 | 10 | 240 | 65 | 32 | 42 |
| **Time Limit**<br>**Imposed for**<br>**Informed Consent** | Yes | No | Yes | No | No | No |
| **Informed Consent**<br>**Mentioned** | Yes | Ongoing | Yes | Yes | Yes | Ongoing |
| **Statement of**<br>**Informed Consent**<br>**Obtained** | Yes | Ongoing | Yes | Yes | Yes | Ongoing |
| **Statement of**<br>**Ethics Approval** | Yes | Ongoing | No | No | Yes | Ongoing |

## RESULTS

Table 1 shows the demographic characteristics of the 6 included trials, 2 of which are ongoing, and 4 of which were published between 2009 and 2016. The trials were conducted collectively in 6 continents and the number of participants in each trial ranged widely from 7 to 3,206 patients. All 4 published trials mentioned the term informed consent and two trials explicitly stated the involvement of an ethics committee in their published manuscript.(32,33)

### Patient Capacity

The median NIHSS score for each completed trial was at least 8, meaning most participants experienced a moderate stroke (Table 1). 5 trials allowed for consent to be obtained from a designated patient proxy if the patient was deemed to lack capacity (Figure 2). Of those 5 trials, 2 required assent from the patient if the patient was consented by proxy and the other two trials did not require patient assent. Trials reporting direct consent (defined as not consent by proxy) of 75% to 100% of their participants were associated non-significantly with shorter DTT times than trials reporting direct consent of 25% to 50% of patients for both the experimental and control arms with a trend towards significance (group = median[IQR]; 75% to 100% experimental = 32[16-48], control = 34[17.5-50.5]; 25% to 50% experimental = 117.5[65-170], control 117.5[65-170]). However, 2 of the 6 trials did impose a time limit for when informed consent needed to take place (Table 1). When trials were asked to comment on challenges that arose during study protocol development, all comments referenced problems delineating specific exclusion or inclusion criteria (Table 2). Additionally, when asked about challenges related to obtaining consent, 3 out of the 5 comments mentioned difficulty with obtaining proxy consent.

**Table 2.** Commentary provided when asked to describe the challenges encountered with study protocol development or in obtaining informed consent from patients.

| **Challenges with Study Protocol Development** | **Challenges in Obtaining Informed Consent** |
| --- | --- |
| We are allowed to include patients in the trial on the basis of their verbal consent. Written informed consent is obtained after treatment has been administered. The rationale behind this is to avoid delay of treatment because of written informed consent. | It is in some situations difficult to obtain a verbal informed consent in acute stroke patients. After the acute phase, it may also be difficult for the patients to give their written informed consent. In our study, it was also be challenging to obtain written informed consent for study participation from patient/proxy if the patient suffers from a complication such as intracranial hemorrhage following treatment. |
| Too numerous to count. This was prior to the era of central IRB's, so each participating institutions had their own IRB weighing in on the consent. While our consent was templated, each participating institution's approved consent form was reviewed by us to be sure it contained all elements required by our IRB and OHRP. | Finding proxies for aphasic patients unable to consent for themselves was sometimes challenging. If no proxy available, the patient could not be enrolled. |
| The study was performed strictly according to SOP for acute stroke. Information and randomized treatment were the only study specific changes. Ethical aspects of informed consent in patients with a brain lesion, even a small one, resulted in lengthy discussions. | We informed the patient verbally, regardless of the patients state. We did not require an oral consent from the patient for the treatment. The patient (or proxy) was later asked for a written consent to use their data for research. |
| Whether patients given endovascular therapy will exclude them from the study. | The safety and efficacy about the study drug. More details on the outcome for the former participants. |

**Figure 2.**
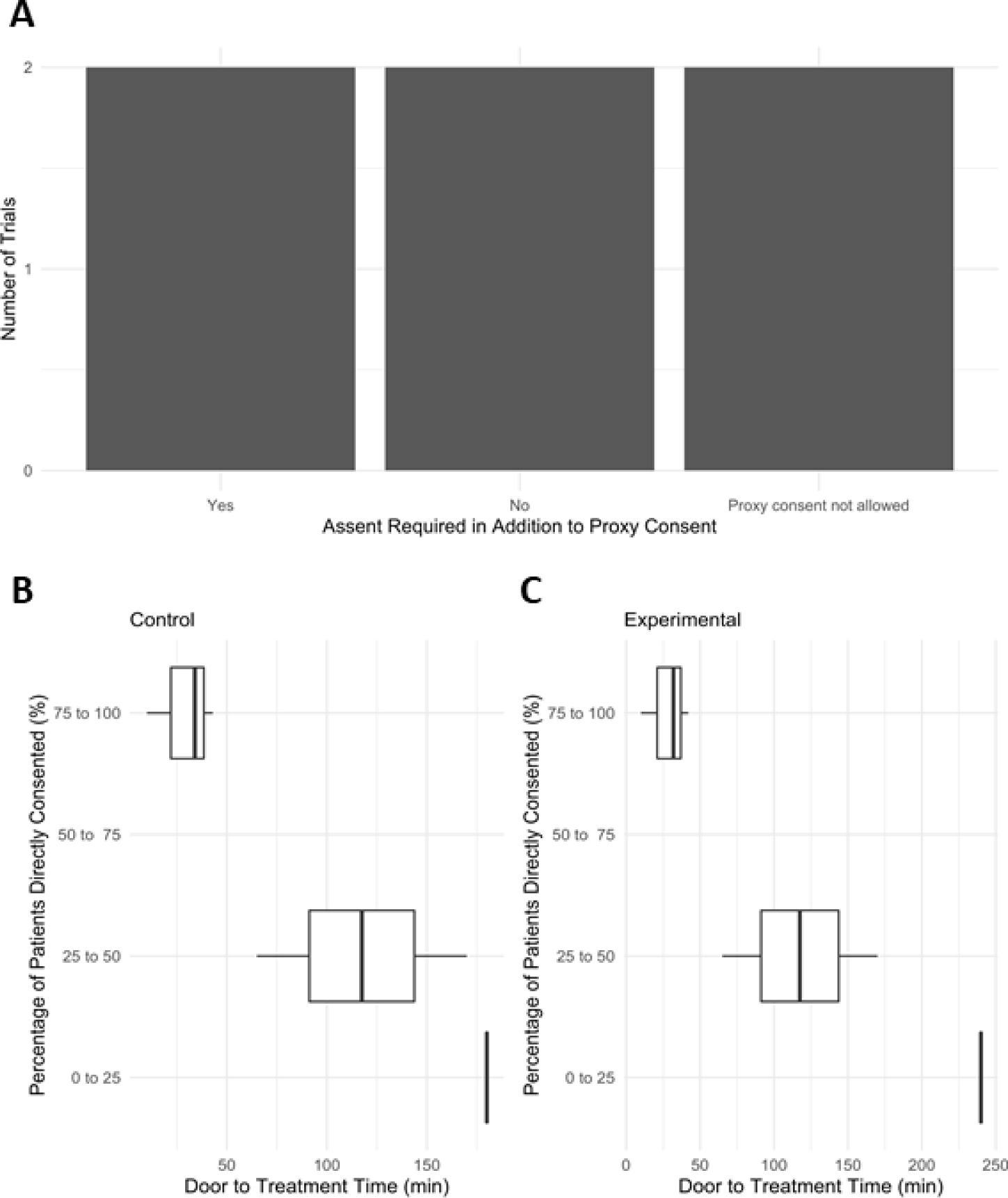
Patients were either directly consented to the trial, consented by proxy with required assent from the patient, or consented by proxy without assent. A) Number of trials requiring assent in addition to consent by proxy. DTT of the B) control arm and C) experimental arm was plotted in relation to percentage of patients directly consented divided into quartiles: 0% to 25% (n=1), 25% to 50% (n=2), 50% to 75% (n=0), 75% to 100% (n=3).

### Logistics of Obtaining Informed Consent

All trials allowed research attendings to obtain informed consent in the ED, but no trial allowed consent to be obtained by nurses, EMTs, undergraduate students, or residents other than a neurology resident (Figure 3). Although some trials allowed consent to take place in the hospital and in 1 case over the phone, no trial allowed for consent to be obtained in an ambulance or while at an outside hospital (OSH) prior to transfer. When asked to describe the required formal training received specific to the informed consent process, only 2 trials mentioned specific training with regards to informed consent skills and 1 trial did not require formal training.

**Figure 3.**
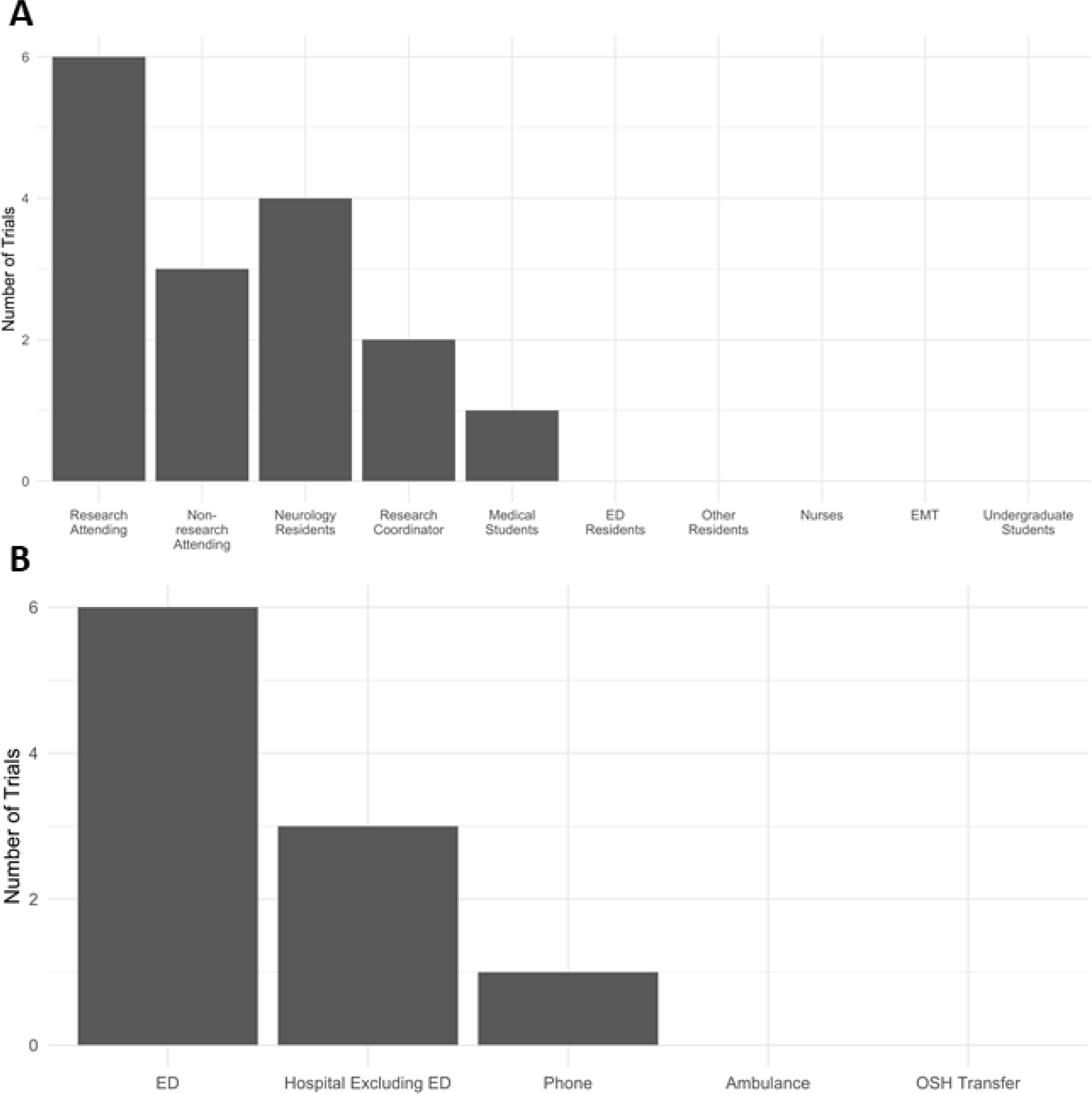
A) The types of professionals obtaining consent and B) the location consent was obtained. The Emergency Department (ED) was considered a separate location from the hospital itself. Outside hospital (OSH) was defined as a hospital other than where the study was being conducted.

### Foreign Language

The 6 trials took place in various continents and 4 out of the 6 trials made translators and translated consent documents available. Interestingly, trials that offered translated consent documents had longer DTT than trials that did not offer translated documents in both the control and the experimental arms (Figure 4). Similarly, trials that offered translation services had longer DTT than trials that did not in both the control and experimental arms.

**Figure 4.**
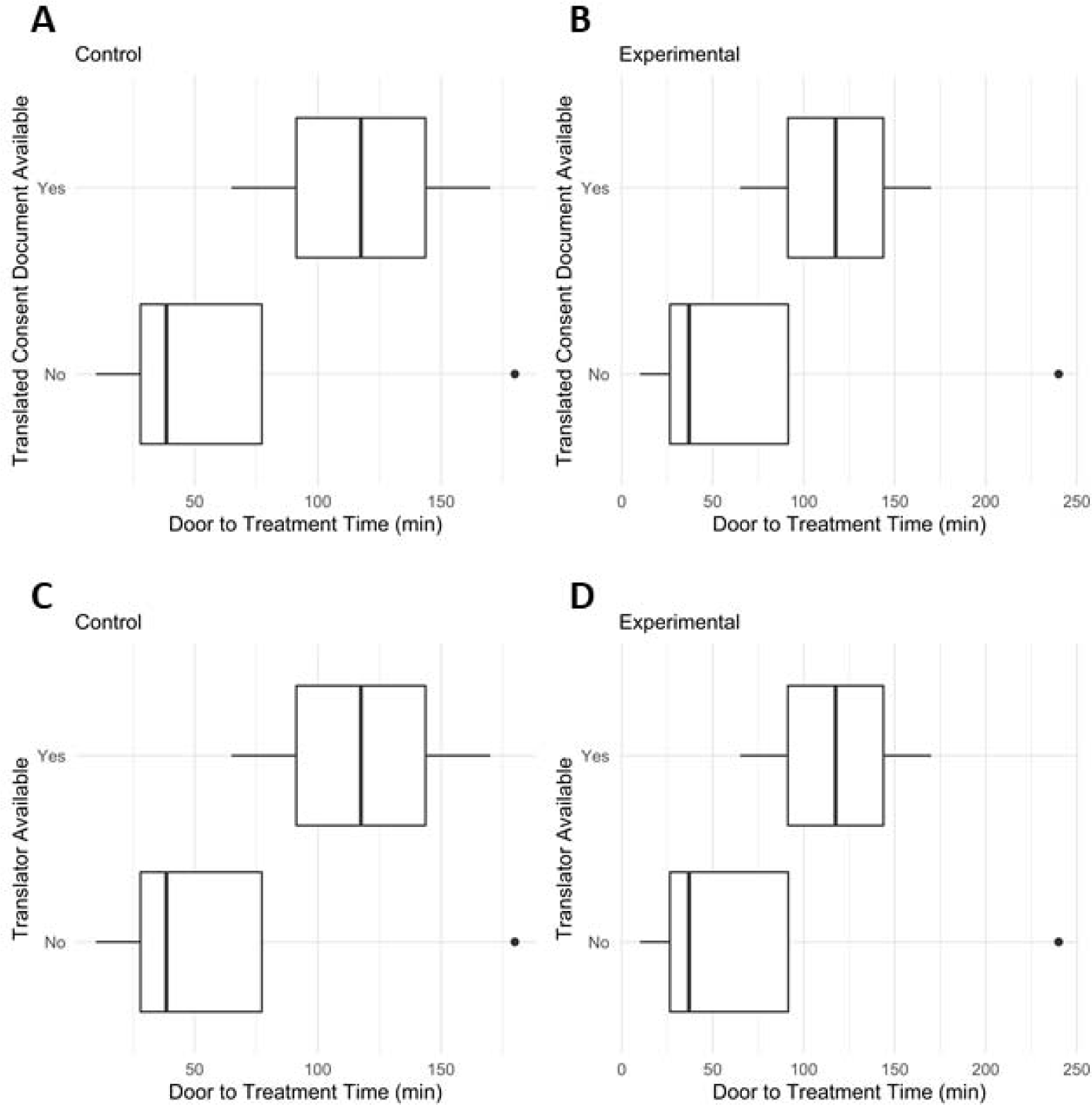
The effect of translator services on the DTT for trials. The DTT for the A+C) control arm and B+D) experimental arm were plotted in relation to having A+B) translated consent documents available (n = 2) or having C+D) a translator available (n=2).

## DISCUSSION

Acute stroke treatment trials face unique challenges in obtaining informed consent due to the time-sensitive nature of acute stroke treatments and a patient population with neurologic deficits. The goal of our study was to characterize the approaches and barriers to obtaining informed consent in trials comparing the effectiveness of tPA with alternative treatment. To our knowledge, this study is the first to investigate the experiences of international stroke trial investigators related to obtaining informed consent.(34) Our findings highlight the many challenges of obtaining consent in the acute setting and suggest that more transparency in current informed consent processes and a willingness to innovate from traditional consent are needed to develop treatments that improve stroke outcomes.

Informed consent is a mechanism for patients to exercise autonomy in trial enrollment, but patients and their surrogates also derive value from informed consent discussions.(35,36) As such, the manner of patient participation is intrinsically an ethical factor in trials without straightforward informed consent processes, such as trials of acute stroke or other emergency treatments. Investigators surveyed from the AbESTT II trial indicated that obtaining informed consent produces unnecessary delays, but simultaneously felt an exemption for informed consent was inappropriate.(37) Thus, the implementation of informed consent presents a conflict between balancing patient-centric care with faster DTT times for superior outcomes, development of superior stroke therapies, and respect for patients as autonomous human beings.(6,37)

### Demographics of Trials

The 6 trials included were conducted at major stroke institutions internationally, with representation from North America, Asia, Europe, Australia, and South America. Because of this variation in location, different institutional and cultural values may have influenced the methods and parameters used for obtaining informed consent, as evidenced in previous studies.(38) One such unique challenge was that a centralized IRB was not yet established before some trials took place (Table 2). Therefore, each participating trial center created an individual consent protocol to be approved by the home institution IRB, which decreased reproducibility of the informed consent process across trial sites. Another significant variation observed was the number of participants: trials ranged from 7 to 3206. It is likely that trials with larger cohorts faced more challenges, but these multi-center trials may have had the motivation and resources to implement more standardized approaches at multiple locations. Although the differences between the trials examined may be related to trial design, legal, institutional, and cultural effects, these trials overall provide evidence of the diverse landscape of informed consent.

### Reporting the Consent Process

Information regarding how consent was obtained, consent forms, timing, and personnel involved across these trials was highly variable, and often could not be located publicly through protocol papers or within the published manuscripts. Interestingly, only half of the trials mentioned the involvement of a research ethics committee in their published results. Ethical oversight in the design of future studies, as is the case in the ongoing AcT QuICR trial, may be a useful approach to mitigate ethical dilemmas and oversee abbreviated methods of informed consent.(26,39) 1 of the 6 trials enrolled patients via a waiver of informed consent. None of the 14 trials made a copy of the informed consent document publicly available. Even after contacting trials for a copy of the informed consent document, only 2 provided forms for further examination. The absence of information highlights the need for research ethics involvement in trial design and greater transparency concerning informed consent in published manuscripts.

### Patient Capacity and Consent by Proxy

Given that patients experiencing a stroke are a vulnerable population on the basis of the neurologic nature of the disease, understanding patient capacity is critical. In this study, 3 of 6 trials reported consenting patients directly for 75 to 100 % of participants, while only one trial reported consenting patients directly for less than 25% of participants (Figure 2). It remains important, however, to include cognitively impaired subjects to determine the generalizability of the proposed interventions.(40) NIHSS scores reported indicate most participants experienced a moderate stroke (Table 1). In the moderate NIHSS group, the ability to cognitively understand circumstances and make an informed decision varies widely.(41,42) This potentially discordant finding between reports that the majority of patients directly consented and had a “moderate” stroke could be due to alternate definitions of direct consent outside of the USA.(43) 4 of the 6 trials in this study were conducted outside of the USA, where deferred consent is commonly practiced.(44,45) Deferred consent involves randomization of the patient into the protocol based on the discretion of the investigator, followed by informed consent during a later phase of care. In that case, investigators may have received assent from the stroke patient in the acute phase and formal written consent from the legally authorized representative. Given this information, there may have been a difference in interpretation of the survey question, where trial representatives responded with a focus on who was actually signing the consent form rather than who initially provided consent before treatment was administered (Supp 1). This may explain why although most patients experienced a moderate stroke, trials responded that over 75% of stroke patients were directly consented. Although deferred consent is one possible solution to ensure patients receive treatment in a timely manner and are appropriately aware of therapies, patient autonomy may become an issue: the investigator cannot take away the medications or treatment already provided, even if the patient were to later decline participation in the trial.

While it is unclear how trials in this study determined capacity for consent, other trials have used aspects of the NIHSS score to assess capacity, including the sections regarding “level of consciousness”, “best gaze”, and “best language.”(15) More research needs to be conducted about ways to best assess capacity in moderate NIHSS patients to determine whether examining aspects of the NIHSS is sufficient to determine capacity.(46) When capacity is lacking, many stroke trials use consent by proxy rather than direct consent to enroll patients.(43) From an ethical perspective, there is concern as to whether surrogates are aligned with the wishes of patients in research trials.(47) To address this issue, patient assent can be required in addition to proxy consent, which was performed in some of the trials in this study.(35) Although surrogate decision makers must make difficult choices, the research team should still involve the patient to their fullest extent by attempting to gain assent when patient capacity is determined to be lacking.

### Logistics of Obtaining Informed Consent

Although communication techniques employed during the consent process between research staff and patients are vital for ethical and efficient obtainment of consent, only a subset of the trials indicated a dedicated training process specific to the trial (Table 2). All trials in this study sample relied on highly educated and experienced providers as well as dedicated research coordinators to obtain consent (Figure 3a). A dedicated training protocol for delivering informed consent can ensure a high standard of communication and create more time to discuss the trial with patients and families.(46,48) A third party involved in obtaining consent may also place less pressure to enroll.(48)

In the trials studied, consent was obtained in a variety of settings; however, only one trial allowed consent to be obtained over the phone and no trials obtained consent upon patient transfer or in an ambulance. Obtaining consent over the phone in a prehospital setting has been performed in the FAST-MAG trial and provided more time for patients to consider consent as well as increased efficiency.(49) We suggest that consent in the pre-hospital setting should be considered for future trials. In addition, 3 out of 5 trials reported challenges obtaining consent by proxy, some of which may be mitigated by phone consent (Table 2). Phone consent is important when no proxy is available within the time window due to geographic constraints.(36,50) However, it can be challenging to reach patient families with distressing information and ask for a decision when they are most vulnerable. Another solution to consider in the case of acute stroke trials may be a waiver of consent, known as an exception from informed consent (EFIC) in the USA, as delays in treatment due to the time it takes to achieve informed consent can lead to worse outcomes for patients.(51,52)

Recent revisions to the Common Rule in the Federal Rules for the Protection of Human Subjects emphasize the responsibility of researchers to deliver information with concise and simple language during the consent process.(53–55) Consent forms in the USA now should emphasize key points for participation up front.(56) This is important because enrollment decisions in stroke trials can occur in a highly stressful environment that may increase cognitive load and diminish comprehension. A third of the trials reviewed implemented a time limit in which consent had to be obtained (Table 1). This approach may be appropriate in the acute setting to indicate the time sensitive nature of the process to research staff and to promote efficiency in the consent process. One trial made note of the decision to allow patients to provide verbal consent to initiate treatment followed by deferred written consent (Table 3). Interpersonal communication may be a more efficient means to obtain consent and produce better understanding in comparison to the medium of paper forms.(48,57) As such, verbal consent has previously been proposed as a function-based approach to informed consent.(58–60) Shortening the consent form is another consideration to reduce complexity, however, a longer consent form may be appropriate for proxy decision making.(35,61,62) Overall, a more targeted consent model may be preferable on an ethical basis due to the time sensitive nature of benefit to the patient and the potential to enhance patient understanding.(17)

**Table 3.** If a trial indicated that professionals obtaining informed consent were required to complete formal training specific to the informed consent process, free text space was given to describe the training process.

| Trial Name | Description of Required Formal Training for Informed Consent |
| --- | --- |
| ENCHANTED | They needed training in the protocol and how to obtain informed consent, specifically related to dealing with equipment. The only people who were allowed to obtain consent were those who had training and listed on a research delegation log approved by the site principal investigator and the project coordinating team. |
| noR-TEST 2 | Not required. |
| IV vs. IA tPA | CITI training. |
| TNKS2B | Computerized learning modules plus face-to-face teaching. |
| noR-TEST | Training course for Local Investigators (LI) by the organizing center. LI providing training in the participating hospitals. |
| TPK Derivative | Training about the guideline for thrombolysis for the acute ischemic stroke. Training about the design of the trial. Training about the skills in conducting informed consent. |

### Foreign Language

Translators and translated consent documents may provide an avenue to greater patient diversity in stroke trials. Trials which included patients with potential language barriers requiring translators and translated consent documents demonstrated a trend towards increased door-to-treatment time, suggesting that efficiency and efficacy may be impacted where such barriers exist (Figure 4). Efforts to increase the availability of rapid bedside interpretation services, such as with interpreter phones and professional translators, may help correct well-documented, ethnicity-based disparities in thrombolysis.(63,64) Additional barriers in the consent process may also exist for patients who speak foreign languages. Patients with limited English proficiency may have lower health literacy, availability of surrogate decision makers, or knowledge of concepts related to study design, such as randomization and clinical equipoise.(65,66) These additional barriers increase the need for rapid translation services and education at the bedside to ensure informed consent is achieved in a timely manner and to promote representation of a diverse patient population.

### Limitations

The findings from this study are limited by the small sample of trials that met inclusion criteria. We also did not have responses from 8 of the 14 trials that met inclusion criteria and could not locate trial protocols or procedure manuals that outlined the informed consent procedures. We welcome any trial investigators or staff to complete our survey through this link http://bit.ly/strokeconsent or by contacting the corresponding author.

Some of the trials were ongoing, and the respondents could not complete all data fields. In cases where the trial was conducted across multiple clinical sites, responses may not reflect site-specific variation in how informed consent was obtained. Trials were additionally limited to studies listed in trial registries in English, thus excluding the pool of international studies which otherwise satisfied inclusion criteria. Besides informed consent, unreported clinical factors, such as blood pressure management, may also influence the DTT times in the included trials.(67)

Finally, the informed consent process for stroke trials may change in the future due to altered interpretation of EFIC.(68) EFIC was drafted in 1996 and finalized in 2013, but has not been frequently implemented in stroke trials.(52,69) Standard of care trials, such as those examined in this study, are typically not candidates for the waiver because of the proven moderate efficacy of tPA.(17,69) Other countries have different regulations and models of consent not captured by this study. A new landscape for informed consent in the acute setting may be necessary to ensure medical care does not stagnate and that research can allow for the development of the next standard of care.

## CONCLUSIONS

Time is most valuable for patients suffering from a stroke: the amount of brain function regained is directly proportional to the time it takes to administer treatment. It is critical that investigators respect this race against time. However, investigators are researching thrombolytic therapies that have the potential to improve the quality of life and provide great benefit to society. Given the regulatory burden and patient family stress, the current informed consent process in the USA does not allow for direct comparison to discover a replacement for tPA. This research cannot progress unless the standards of informed consent change or waivers are permitted, but we must balance protecting the individuals whose willing involvement enables such pivotal discoveries. Therefore, we recommend a new era of informed consent in acute stroke trials with ethical oversight to develop a targeted approach to informed consent based on European and Asian models, where most patients are directly consented. We acknowledge that uniform standards of obtaining consent may not be applicable in all trials or scenarios, but future trials should be designed with an emphasis on communication with patients of diverse backgrounds, robust consent protocols, and transparency in the informed consent process. This new approach will pave the way for more streamlined and inclusive study of treatments in acute stroke.

## Data Availability

Data that were used in this study are available on request from the corresponding author.

## ACKNOWLEDGEMENTS

This research was unfunded. The authors wish to thank participating studies; without their assistance, this research would not have been possible. Authors also wish to thank James F. Burke MD, MS for his critical editing of this manuscript and Lesli E. Skolarus, MD for her expert review of the survey.

