## Supplementary Figure 1 for "Ethical Considerations during the Informed Consent Process for Acute Ischemic Stroke in International Clinical Trials"

**SUPPLEMENTARY FIGURE 1.** A complete copy of the survey administered to stroke trials.

**Ethical Considerations of Informed Consent in the Setting of Acute Ischemic Stroke.** Thank you for taking this brief survey that can be completed in under 10 minutes. This survey was created by a research group at the University of Michigan to assess how informed consent was obtained in a population that may have challenges with standard consent due to lacking capacity in the setting of acute stroke and language or motor deficits that would make it difficult to provide verbal or written consent. Your participation in this research study is voluntary. If you decide to participate in this research survey, you may withdraw at any time. All data is stored in a password protected electronic format. The results of this study will be used for scholarly purposes only. This research study was reviewed by the University of Michigan Institutional Review Board for Human Subjects Research and determined to be exempt under IRB #HUM00180410. If you have more questions concerning this study, please feel free to contact us at.

Q1 Please provide the name of your trial or study.

Q3 What continent or continents was your trial conducted in? Select all that apply.

▢	Africa (32)

▢	Asia (34)

▢	Australia (35)

▢	Europe (36)

▢	North America (37)

▢	South America (38)

Q2 Is your trial currently ongoing?

o Yes (1)

o No (2)

Q4 Please provide or estimate the median door to treatment time in your trial for the CONTROL group in minutes?

________________________________________________________________

Q29 Please provide or estimate the median door to treatment time in your trial for ALL EXPERIMENTAL groups in minutes?

________________________________________________________________

Q5 Was the above information estimated or calculated from data?

o Estimated (1)

o Calculated (2)

Q6 Was a translator used to obtain informed consent if the patient did not speak the primary language of your country?

o Yes (1)

o No (2)

o Unknown (3)

Q7 Were versions of the informed consent document available in different languages for patients?

o Yes (1)

o No (2)

o Unknown (3)

Q8 What types of professionals were allowed to obtain informed consent to participate in the trial? Choose all that apply.

▢	research attending physician (1)

▢	non-researcher attending physician (2)

▢	ED residents (3)

▢	Neurology residents (4)

▢	Other residents (5)

▢	Nurses (6)

▢	EMT (Emergency Medical Technicians) (7)

▢	Research coordinator (8)

▢	Medical students (9)

▢	Undergraduate students (10)

▢	Other (11)

Q9 Did the professionals obtaining informed consent complete required formal training specific to the informed consent process for this trial?

o Yes (1)

o No (2)

o Unknown (4)

Skip To: Q11 If Did the professionals obtaining informed consent complete required formal training specific to th... = No

Q10 If the professionals conducting informed consent completed required formal training, please describe the process here.

________________________________________________________________

________________________________________________________________

________________________________________________________________

________________________________________________________________

________________________________________________________________

Q11 What modalities were used to present the informed consent to patients and their proxies? Choose all that apply.

▢	Written document (1)

▢	Verbal discussion (2)

▢	Video (3)

▢	Other (5) ________________________________________________

Q12 What modalities were used to document the informed consent process from patients and their proxies? Choose all that apply.

▢	Written signature (1)

▢	Electronic signature (2)

▢	Verbal affirmation (3)

Q13

In what settings could informed consent be obtained? Choose all that apply.

▢	Hospital (not Emergency Department) (1)

▢	Emergency Department (2)

▢	Ambulance (3)

▢	An outside hospital prior to transfer (4)

▢	Over the phone (5)

Q14 Was there a time limit imposed for informed consent to take place, (i.e. within 60 minutes of arrival to the hospital)?Edit Question Label

o Yes (1)

o No (2)

o Unknown (3)

Skip To: Q16 Was there a time limit imposed for informed consent to take place, (i.e. within 60 minutes of arr... = No

Skip To: Q16 Was there a time limit imposed for informed consent to take place, (i.e. within 60 minutes of arr... = Unknown

Q15 If there was a time limit imposed for informed consent to take place, what was the time limit in minutes?

________________________________________________________________

Q16 What percentage of the time was the patient able to provide informed consent VS a proxy/a waiver from informed consent?

o 0% to	(1)

o 25% to	(2)

o 50% to	(3)

o 75% to 100% (4)

o unknown (5)

Q17 Was an attempt made to contact a proxy for trial participation?

o Yes (1)

o No (2)

o Unknown (3)

Q18 If the patient was able to assent, was this required in addition to consent by proxy?

o Yes (1)

o No (2)

o Does not apply (3)

Q19 Were there any cases in which informed consent was waived due to the need for emergency care but the patient was still enrolled in the trial?

o Yes (1)

o No (2)

o Unknown (3)

Q22 If applicable, please comment on any challenges encountered with study protocol development.

________________________________________________________________

________________________________________________________________

________________________________________________________________

________________________________________________________________

________________________________________________________________

Q23 If applicable, please comment on any challenges that you or your team encountered in obtaining informed consent from patients.

________________________________________________________________

________________________________________________________________

________________________________________________________________

________________________________________________________________

________________________________________________________________

Q24 If you would be willing to be recontacted in case there are follow up questions, please provide your email below.

________________________________________________________________
